# ANTsX neuroimaging-derived structural phenotypes of UK Biobank

**DOI:** 10.1101/2023.01.17.23284693

**Authors:** Nicholas J. Tustison, Michael A. Yassa, Batool Rizvi, Philip A. Cook, Andrew J. Holbrook, Mithra T. Sathishkumar, Mia G. Tustison, James C. Gee, James R. Stone, Brian B. Avants

## Abstract

UK Biobank is a large-scale epidemiological resource for investigating prospective correlations between various lifestyle, environmental, and genetic factors with health and disease progression. In addition to individual subject information obtained through surveys and physical examinations, a comprehensive neuroimaging battery consisting of multiple modalities provides imaging-derived phenotypes (IDPs) that can serve as biomarkers in neuroscience research. In this study, we augment the existing set of UK Biobank neuroimaging structural IDPs, obtained from well-established software libraries such as FSL and FreeSurfer, with related measurements acquired through the Advanced Normalization Tools Ecosystem. This includes previously established cortical and subcortical measurements defined, in part, based on the Desikan-Killiany-Tourville atlas. Also included are morphological measurements from two recent developments: medial temporal lobe parcellation of hippocampal and extra-hippocampal regions in addition to cerebellum parcellation and thickness based on the Shanneman anatomical labeling. Through predictive modeling, we assess the clinical utility of these IDP measurements, individually and in combination, using commonly studied phenotypic correlates including age, fluid intelligence, numeric memory, and several other sociodemographic variables. The predictive accuracy of these IDP-based models, in terms of root-mean-squared-error or area-under-the-curve for continuous and categorical variables, respectively, provides comparative insights between software libraries as well as potential clinical interpretability. Results demonstrate varied performance between package-based IDP sets and their combination, emphasizing the need for careful consideration in their selection and utilization.

## 1 Introduction

UK Biobank (UKBB) is a unique epidemiological effort which aims to prospectively identify potential relationships between disease and associated risk factors through the leveraging of comprehensive individualized medical and sociodemographic data. Enrollment began in 2006 and continued for four years ultimately resulting in a cohort of approximately 500,000 individuals. Volunteer age criteria was limited to birth years between 1934 and 1971—an optimal range for observing the onset of certain diseases and their subsequent progression. Continued monitoring of a significant subset is expected to continue for at least 30 years facilitated, in part, by coordination with the National Health Services of the UK. This has resulted in several studies exploring a wide variety of research topics (e.g., the relationship between age and cognitive decline,^1^ the association of polygenic profiles and mental health,^2^ and potential collider bias in COVID-19 assessment).^3^

An integral component of UKBB is the subset of approximately 50,000 subjects who underwent comprehensive imaging batteries, including neuroimaging,^4,5^ specifically structural T1-weighted MPRAGE and T2-FLAIR MRI; diffusion-weighted MRI; resting-state and task-based functional MRI; and susceptibility-weighted MRI. Employing specialized processing pipelines, these raw imaging data are used to generate various quantities, referred to as image-derived phenotypes (IDPs), for use as potential biomarkers. A sampling of resulting image-based research studies evinces insights into such topics as hippocampal volumetric nomograms across age;^6^ population modeling of age, fluid intelligence, and neuroticism;^7^ and brain structural changes associated with COVID-19 and the corresponding cognitive effects.^8^

Facilitating the majority of existing UKBB imaging-related research is the FMRIB Software Library (FSL)^9^ which has been specifically tailored to provide UKBB IDPs.^4,5^ For the structural data alone, this includes global and cortical IDPs from FMRIB’s Automated Segmentation Tool (FAST),^10^ subcortical IDPs from FMRIB’s Integrated Registration and Segmentation Tool (FIRST),^11^ and white matter hyperintensity (WMH) load using the Brain Intensity AbNormality Classification Algorithm (BIANCA).^12^ UKBB was subsequently augmented with FreeSurfer-based IDPs^13^ which include both the standard “aseg” segmentation, hippocampal subfield,^14^ and amygdala nuclei^15^ pipeline outputs.

Analogously, the Advanced Normalization Tools Ecosystem (ANTsX) is a collection of interrelated, open-source software libraries for biological and medical image processing and analysis^16^ with developmental roots in high-performing medical image registration^17,18^ and built on the Insight Toolkit (ITK).^19^ ANTsX-based IDPs have demonstrated utility in several studies spanning a variety of organ systems, species, and imaging modalities (e.g.,).^20–22^ These IDPs include those which have been previously reported, such as global brain tissue volumes^23^ and more localized, FreeSurfer-analogous cortical thickness values^16,24,25^ averaged over the Desikan-Killiany-Tourville (DKT) regions.^26^ In addition, recently developed ANTsX functionality includes a medial temporal lobe (MTL) parcellation framework known as “DeepFLASH,” a neural network for segmenting hippocampal subfields and extrahippocampal regions which extends previous work.^27^ Newly introduced functionality also includes regional cerebellum measurements based on the Schmahmann atlas^28^ including cortical thickness (cf).^29^

Characterizing the respective sets of FSL, FreeSurfer, and ANTsX IDPs and their mutual relationships can guide researchers in their usage as there are both significant overlap and notable differences between these measures. And although comparison between sets is potentially insightful, a focused, package-wise comparison using UKBB is difficult due to 1) the absence of complete, individual IDP correspondence across packages and 2) the general purpose of UKBB data (in contrast, for example, to the ADNI data^30^ set which focuses on Alzheimer’s disease). Regarding IDP differences, even between identically defined IDPs (e.g., hippocampal volume), *observer bias*^1^ is a possible source of measurement variance.^31^ Note that this variance is not indicative of inaccuracy, per se, such as with *instrumentation bias* where sub-optimal calibration of software is used as a straw-man for comparative purposes.^32^ Rather, observer bias is supplemental to conventional signal noise considerations as a potential source of measurement discrepancy which can provide insight when considered within the appropriate context. For example, differing labeling protocols for specific anatomical structures, such as the hippocampal subfields and parahippocampal subregions, can reveal differences and those differences can motivate and facilitate harmonization.^33^

To this end, in addition to the core contribution of providing ANTsX-based UKBB IDPs, we explore the similarities and differences between the respective sets of structural IDPs and their combination. Given the usage and availability of several powerful machine learning (ML) techniques, we quantify performance using multiple predictive approaches for tabulated IDP data including gradient boosted decision trees (i.e., XGBoost)^34^ and tailored deep learning networks (specifically, TabNet).^35^ Such frameworks potentially have the additional advantage of providing clinical interpretability of individual features (e.g., SHAP).^36^ For example, one of the most well-studied neuroimaging structural correlative relationships is chronological and so-called “brain age” and their health-dependent divergence.^37^ Such subject-specific, single values are estimated using a variety of ML approaches and IDPs. Although establishing normative values over the human life span has clinical utility, as pointed out in Nyberg,^38^ the single-valued brain age is at the extreme end of an “optimal balance between integration and diversification” required for neuroimaging studies. A single score or index most likely does not capture the extent of the non-linearity and heterogeneity of age and other effects on brain structure.^39^ In contrast, the type of feature-based investigation performed here reveals insight into such questions as: “In what ways do the different IDP sets perform in terms of their predictive capabilities?,” “How does this performance vary with different sociodemographic variables?,” “In what ways are features complementary and can their combined effect improve prediction performance?,” and “How does this performance vary with ML technique?”.

## 2 Materials and Methods

### 2.1 UK Biobank data description

The study was conducted under UKBB Resource Application ID 63965. The total number of subjects at the time of download was 502,413 with 49,351 subjects having undergone the standard imaging battery. Of these imaging subjects, only 40,898 complete sets of downloaded IDPs were in common between those provided by FSL and FreeSurfer processing streams.^5^ Intersection with the final ANTs processed set resulted in a total study cohort size of 40,796.

### 2.2 FSL structural phenotypes

All structural FSL IDPs were included for consideration.^40^ These included the following categories:

- FAST regional grey matter volumes (Category ID: 1101);
- FIRST subcortical volumes (Category ID: 1102);
- global brain tissue volumes and related quantities (Field ID: 25000–25010, 25025); and
- total volume of WMH load (Field ID: 25781)

for a total of 139*_F AST_* + 14*_F IRST_* + 12*_Global_*+ 1*_W MH_*= 166 IDPs.

### 2.3 FreeSurfer structural phenotypes

Several categories of IDPs are available for FreeSurfer comprising a total of 1242 measurements.^40^ However, to make the study dataset more computationally tractable and reduce set size differences between packages, we selected the following popular IDP subsets:

- ASEG volumetric measurements (Category ID: 190);
- DKT volumes and mean thicknesses (Category ID: 196); and
- hippocampal subfields and amygdala nuclei (Category ID: 191) totaling 56*_ASEG_* + 124*_DKT_* + 121*_hipp_* = 301 individual IDPs.

### 2.4 ANTsX structural phenotypes

Both sociodemographic and bulk image data were downloaded to the high performance cluster at the University of Virginia for processing. Grad-warped distortion corrected^41^ T1weighted and FLAIR image data were used to produce the following ANTsX IDPs:

- Deep Atropos brain tissue volumes (i.e., CSF, gray matter, white matter, deep gray matter, brain stem, and cerebellum);
- DKT DiReCT cortical thickness and volumes;
- DKT-based regional volumes;
- DeepFLASH regional volumes;
- Cerebellum regional thickness and volumes;
- Regional WMH loads

totaling 7*_DeepAtropos_* + 88*_DKT reg_* + 128*_DKT DiReCT_* + 20*_DeepF LASH_* + 48*­* + 13*_W MH_* = 302 IDPs which are illustrated in Figure 1. We have reported previously on the first three categories of ANTsX IDPs^16^ but provide a brief description below. We also provide further details concerning both DeepFLASH and the ANTsXNet-ported WMH algorithms.

**Figure 1:**
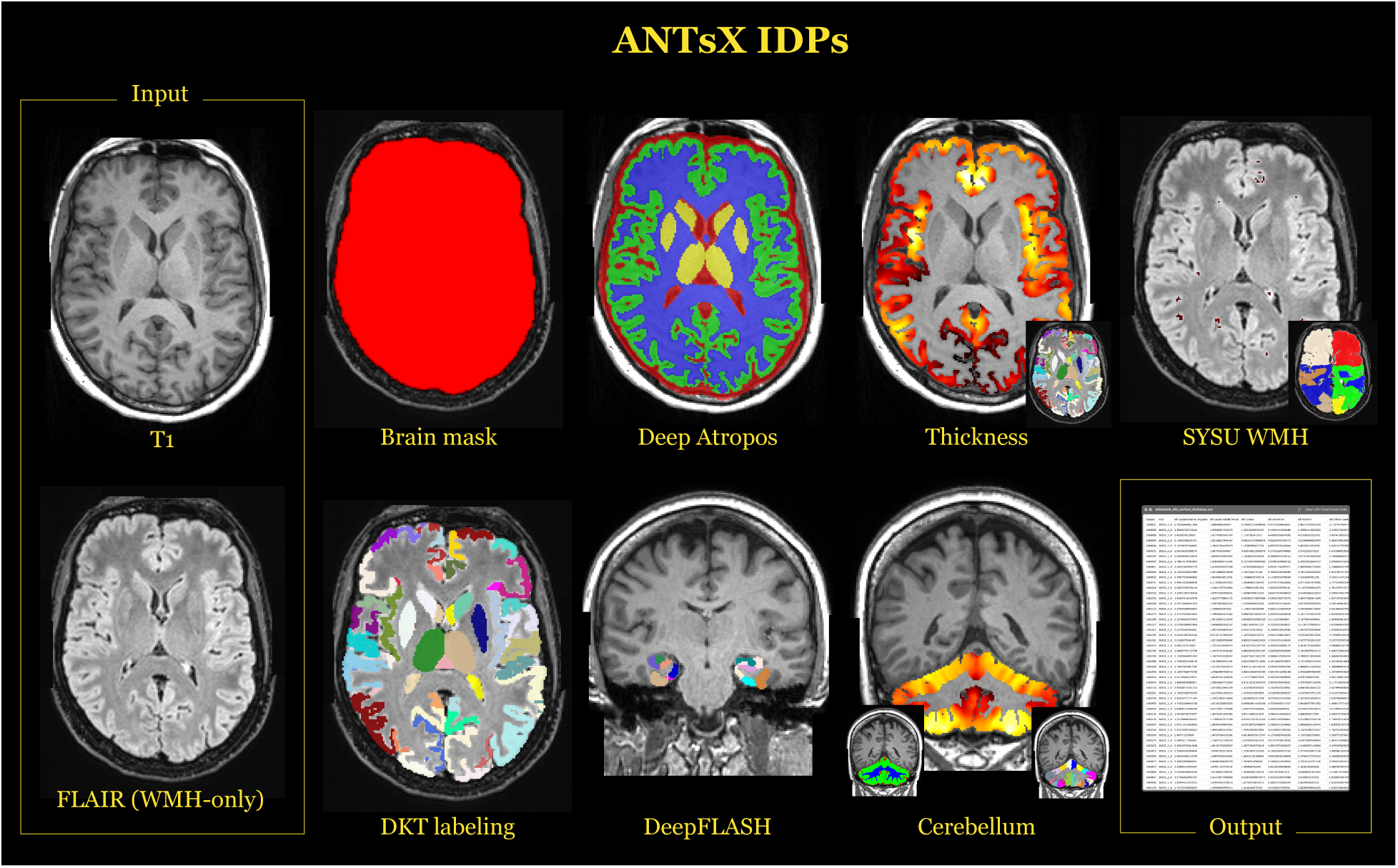
Illustration of the IDPs generated with ANTsX ecosystem tools. Using the gradient-distortion corrected versions of the T1 and FLAIR images, several categories of IDPs were tabulated. These include global brain and tissue volumes, cortical thicknesses averaged over the 62 DKT regions, WMH intensity load per lobe based on the SYSU algorithm, cortical and subcortical volumes from the DKT labeling, MTL regional volumes using DeepFLASH, and morphological cerebellum quantities.

#### 2.4.1 Brain tissue volumes

The ANTsXNet deep learning libraries for Python and R (ANTsPyNet and ANTsRNet, respectively) were recently described in^16^ where they were evaluated in terms of multi-site cortical thickness estimation. This extends previous work^24,25^ in replacing key pipeline components with deep learning variants. For example, a trained network, denoted *Deep Atropos*, replaced the original Atropos algorithm^23^ for six-tissue segmentation (CSF, gray matter, white matter, deep gray matter, cerebellum, and brain stem) similar to functionality for whole brain deep learning-based brain extraction.

#### 2.4.2 DKT cortical thickness, regional volumes, and lobar parcellation

As part of the deep learning refactoring of the cortical thickness pipeline mentioned in the previous section, a framework was developed to generate DKT cortical and subcortical regional labels from T1-weighted MRI.^16^ This facilitates regional averaging of cortical thickness values over that atlas parcellation as well as being the source of other potentially useful geometry-based IDPs. In terms of network training and development, using multi-site data from,^24^ two separate U-net^42^ networks were trained for the “inner” (e.g., subcortical, cerebellar) labels and the “outer” cortical labels, respectively. Similar to Deep Atropos, preprocessing includes brain extraction and affine transformation to the space of the MNI152 template^43^ which includes corresponding prior probability maps. These maps are used as separate input channels for both training and prediction—a type of surrogate for network attention gating.^44^ Using FreeSurfer’s DKT atlas label-to-lobe mapping,^45^ we use a fast marching approach^46^ to produce left/right parcellations of the frontal, temporal, parietal, and occipital lobes, as well as left/right divisions of the brain stem and cerebellum. Using the segmentation output from Deep Atropos, the DiReCT algorithm^29^ generates the subject-specific cortical thickness map which, as previously mentioned, is summarized in terms of IDPs by DKT regional definitions. Given the diffeomorphic and thickness constraints dictated by the DiReCT algorithm, we generate additional DKT regional labels (cortex only) from the non-zero cortical thickness regions to also be used as IDPs.

#### 2.4.3 Fused labeling for automated segmentation of the hippocampus and extrahippocampal regions (DeepFLASH)

A set of IDPs was generated using a deep learning-based framework for hippocampal and extra-hippocampal subfield parcellation which is also publicly available within ANTsXNet (refered to as “DeepFLASH”). This work constitutes an extension of earlier work,^27^ based on joint label fusion (JLF),^47^ which has been used in a variety of studies.^48–53^ DeepFLASH comprises both T1/T2 multi-modality and T1-only imaging networks for parcellating the following MTL regions (cf Figure 2):

**Figure 2:**
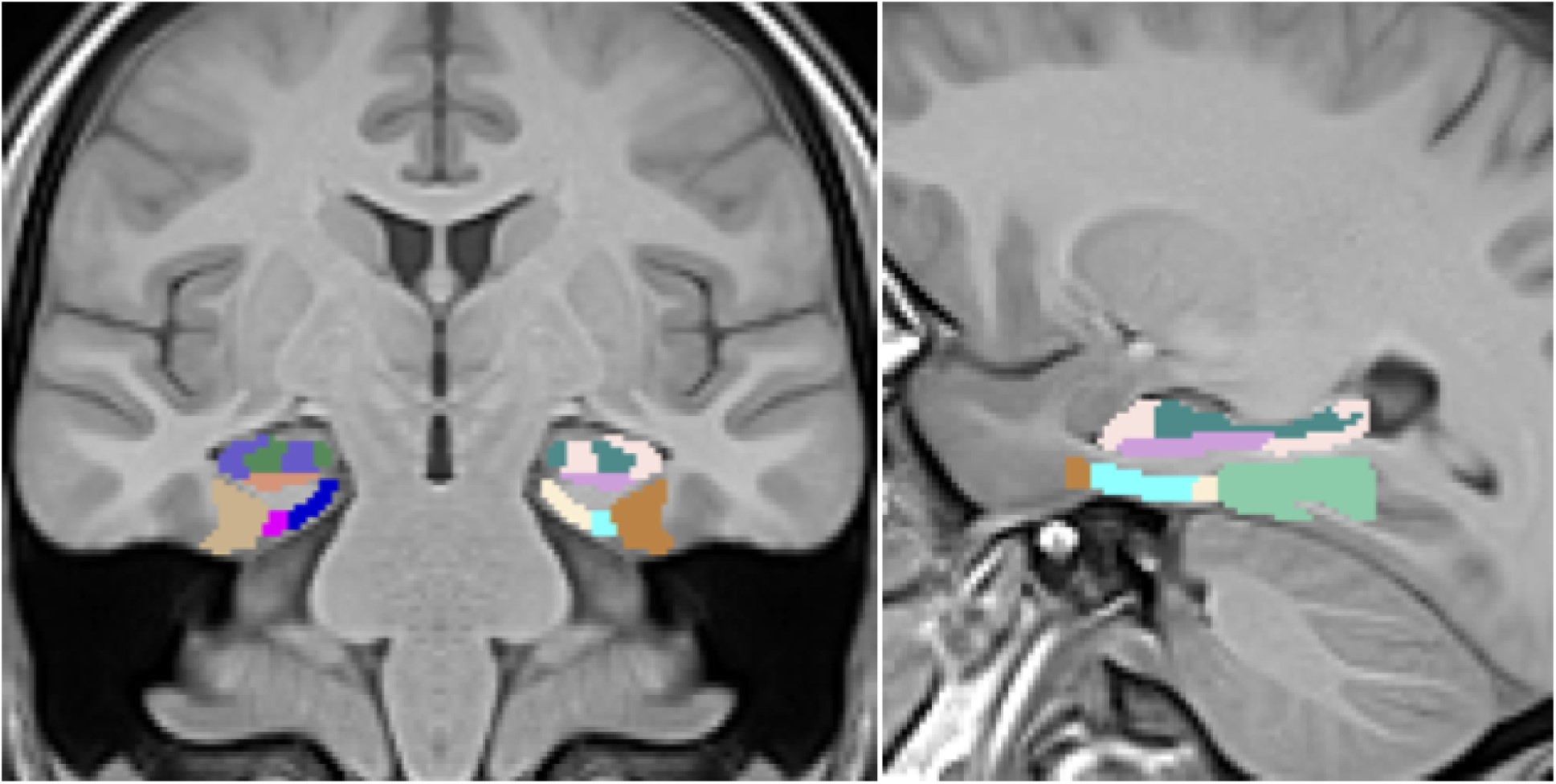
Coronal (left) and sagittal (right) views of the MTL parcellation generated from DeepFLASH superimposed on the T1 template (T2 not shown) used for prediction. The template pose is oriented analogously to hippocampal specific MR acquisition protocols for a tailored segmentation domain.

**Figure 3:**
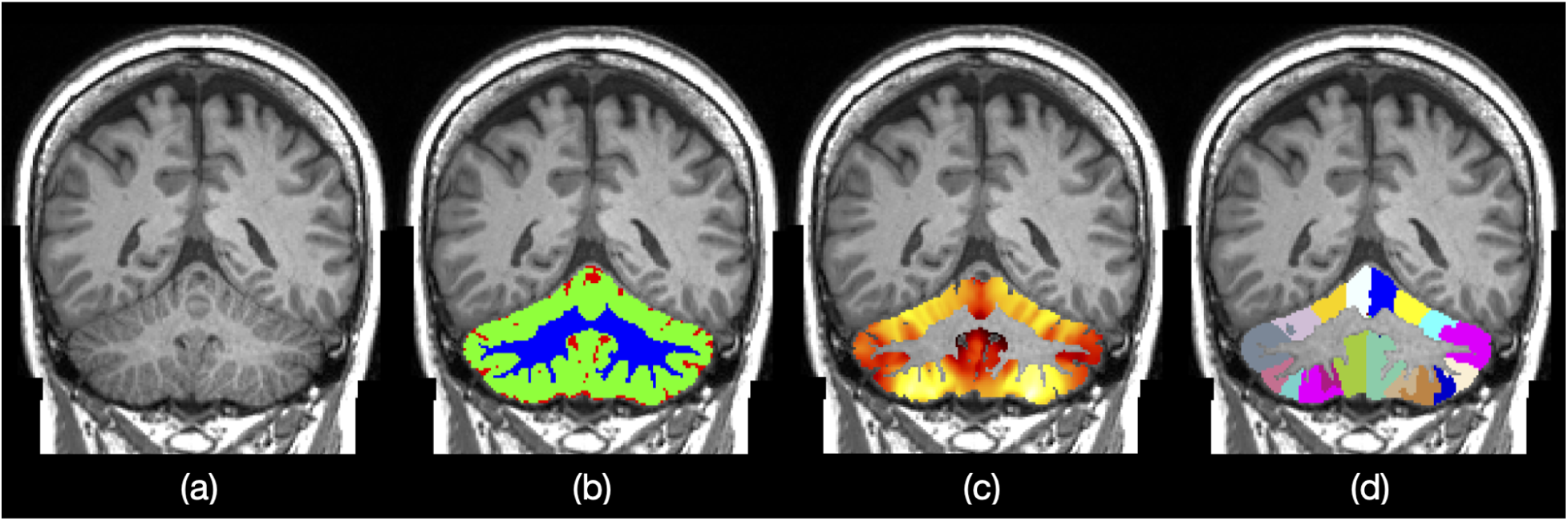
ANTsX cerebellum regional measurements. (b) Three-tissue segmentation (i.e., CSF, gray matter, white matter). (c) Voxelwise, cortical thickness maps. (d) Regional labels based on the Schmahmann atlas.

- hippocampal subfields
  **–** Dentate gyrus/cornu ammonis 2–4 (DG/CA2/CA3/CA4)
  **–** cornu ammonis 1 (CA1)
  **–** subiculum

- extra-hippocampal regions
  **–** perirhinal
  **–** parahippocampal
  **–** antero-lateral entorhinal cortex (aLEC)
  **–** posteromedial entorhinal cortex (pMEC)

DeepFLASH employs a traditional 3-D U-net model^42^ consisting of five layers with 32, 64, 96, 128, and 256 filters, respectively. In addition to the multi-region output, three additional binary outputs (the entire medial temporal lobe complex, the whole hippocampus, and the extra-hippocampal cortex) are incorporated as a hierarchical structural output set. Data augmentation employed both randomized shape (i.e., linear and deformable geometric perturbations) and intensity variations (i.e., simulated bias fields, added noise, and intensity histogram warping). Further information regarding training and prediction can be found at our ANTxNet GitHub repositories.^54,55^

#### 2.4.4 Cerebellum morphology

ANTsX cerebellum IDPs comprise both regional volumes and cortical thickness averages based on the Schmahmann atlas^28^ for cerebellar cortical parcellation. Cortical regions include the following left and right hemispherical lobules:

- I/II
- III
- IV
- V
- VI
- Crus I
- Crus II
- VIIB
- VIIIA
- VIIIB
- IX
- X

Quantifying cerebellar cortical thickness utilizes the DiReCT algorithm.^29^ Both tissue segmentation (CSF, gray matter, and white matter) and regional parcellation is based on a similar deep learning network as that described previously for DeepFLASH. Training data^56^ was coupled with previously described data augmentation. In contrast to DeepFLASH which utilized a single network with multiple outputs, cerebellum output is deribed from first extracting the whole cerebellum and then using it as input to both the tissue segmentation network and Schmahmann regional atlas network.

#### 2.4.5 White matter hyperintensity segmentation

Although UKBB includes white matter hyperintensity segmentation masks^5^ derived from FMRIB’s BIANCA tool,^12^ a recently developed WMH segmentation framework from the “SYSU” team^57^ was imported into the ANTsXNet libraries for WMH segmentation. As discussed in,^58^ this was the top performing algorithm at the International Conference on Medical Image Computing and Computer Assisted Intervention (MICCAI) held in 2017. Image data from five sites were used for both training and testing of segmentation algorithms from 20 different teams. Both the architecture and ensemble weights were made publicly available by the SYSU team which permitted a direct porting into ANTsXNet.

#### 2.4.6 Implementation

Implementations of the previously described pipelines are available in Python and R through our respective ANTsPy/ANTsPyNet and ANTsR/ANTsRNet libraries hosted in the ANTsX ecosystem on GitHub (http://www.github.com/ANTsX/). The specific functions are as follows:

- ANTsPyNet (Python)
  **–** brain_extraction(…)
  **–** deep_atropos(…)
  **–** cortical_thickness(…)
  **–** desikany_killiany_tourville_labeling(…)
  **–** deep_flash(…)
  **–** cerebellum_morphology(…)
  **–** sysu_media_white_matter_segmentation(…)

- ANTsRNet (R)
  **–** brainExtraction(…)
  **–** deepAtropos(…)
  **–** corticalThickness(…)
  **–** desikanyKillianyTourvilleLabeling(…)
  **–** deepFlash(…)
  **–** cerebellumMorphology(…)
  **–** sysuMediaWhiteMatterSegmentation(…)

Self-contained examples of all listed functionality are available as part of an online ANTsX tutorial also hosted on GitHub.

### 2.5 Predictive modeling for IDP characterization

Insight into the relationships between neurostructural and phenotypic measures is often possible through predictive modeling of sociodemographic targets and neuroimaging biomarkers. Many strategies for data exploration leverage standardized quantities derived from existing pipelines, which constitutes a form of dimensionality reduction or feature extraction based on clinically established relevance. Such tabulated data has several advantages over direct image use including being relatively easier to access, store, and manage. Analyses with off-the-shelf statistical packages is also greatly simplified. Additionally, using standardized features in predictive modeling, where feature importance is a component of the analysis, significantly facilitates the clinical interpretability of the modeling process.

Herein, we compare predictive modeling frameworks using tabular data. Baseline comparisons include standard linear regression where linear dependencies between covariates were resolved using findLinearCombos of the caret R package.^59^ Although the number of observations relative to the number of covariates for the specified models is sufficiently large, given the constraints of traditionally-sized data sets, we also compared results based on sparse linear regression, specifically, the Lasso method as found in the glmnet R package,^60^ using recommended parameters.

We also evaluated two popular packages for use with tabular data, viz. XGBoost^34^ and TabNet.^35^ The former is a popular implementation of gradient boosted decision trees known for superb performance and computational efficiency. Similar to random forests,^61^ gradient boosted decision trees leverage ensembles of weak classifiers (i.e., individual decision trees) to enhance model accuracy. However, in contrast to random forests, which generate weak classifiers using random initialization, gradient boosted decision trees are constructed in stages based on the gradient of a specified cost function.^62^ The following common hyperparameters were used: maximum tree depth = 6, number of rounds = 1000 (with early stopping after 10 rounds of no improvement), squared error as the loss function for regression targets, and logistic and softmax losses for binary and multi-label classification problems, respectively.

TabNet is a deep learning framework specifically engineered for structured tabulated data which incorporates sparsity considerations as well as providing feature importance for interpretability. We used an established PyTorch implementation of TabNet^63^ with the default parameters which is reported to demonstrate good performance on a variety of predictive problem types. While deep learning methods have proven both highly popular and effective within neuroimaging research, much of this work has been restricted to convolutional neural networks for image-based analyses, as opposed to parallel research with tabular data, hence the motivation for the development of TabNet. Much more generally, this is a current topic of interest for the larger machine learning community.^64–66^ As a final baseline comparison, we used a basic dense neural network (DenseNet) consisting of two hidden layers with 512 units in the first layer and 256 units in the second layer using leaky ReLU activation where the output layer employed linear activation for regression and sigmoid/softmax for binary/multi classification.

For each of these predictive modeling frameworks, we selected several target variables for our comparative evaluation (cf. Table 1) and generated models of the form:

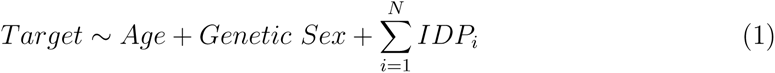

where *i* indexes over the set of *N* IDPs for a particular grouping. In the cases where *Age* or *Genetic Sex* is the target variable, it is omitted from the right side of the modeling equation.

**Table 1:** Set of UKBB sociodemographic targets for evaluation.

| Target | Data ID | Brief Description |
| --- | --- | --- |
| Age | 21003-2.0 | Age (years) at imaging visit |
| Fluid intelligence score | 20191-0.0 | Number of correct answers (of 13) |
| Neuroticism score | 20127-0.0 | Summary of 12 behaviour domains |
| Numeric memory | 20240-0.0 | Maximum digits remembered correctly |
| Body mass index | 21001-2.0 | Impedance-based body composition |
| Townsend deprivation index | 189-0.0 | Material deprivation measure |
| Genetic sex | 22001-0.0 | Sex from genotyping |
| Hearing difficulty | 2247-2.0 | Any difficulty? Yes/No |
| Risk taking | 2040-2.0 | Do you take risks? Yes/No |
| Same sex intercourse | 2159-2.0 | Ever had? Yes/No |
| Smoking | 1249-2.0 | Daily, occasionally, or $\leq 2$ times? |
| Alcohol | 1558-2.0 | Six frequency categories<sup>†</sup> |

†: Daily, 3-4 times/week, 1-2 times/week, 1-3 times/month, special occasions only, and never.

Assessment of the model groups based on the three individual sets of IDPs and their combination employs standard quality measures: area under the curve (AUC) for classification targets and root-mean-square error (RMSE) for regression targets. We also explored individual IDP importance through the use of model-specific parameter assessment metrics such as the absolute value of the t-statistic for linear models and SHAP values^36^ for neural networks.

## 3 Results

### 3.1 Package-wise Group IDP Comparison

To compare the groups of IDPs, we used the three IDP sets (FSL, FreeSurfer, ANTsX) and their combination (“All”) to train predictive models using the preselected target sociodemographic variables from Table 1. We first revisit a previous evaluative framework of ANTsX cortical thickness values by comparing their ability to predict *Age* and *Genetic Sex* with corresponding FreeSurfer cortical thickness values.^16^

Following this initial comparative analysis, ten-fold cross validation, using random training/evaluation sampling sets (90% training/10% evaluation), per IDP set per target variable per machine learning technique (i.e., linear regression, Lasso, XGBoost, DenseNet, and TabNet) was used to train and evaluate the models described by Equation (1).

#### 3.1.1 Revisiting ANTs and FreeSurfer cortical thickness comparison

In,^16,24^ IDPs under consideration were limited to ANTsX-based and FreeSurfer cortical thickness measurements averaged over the 62 regions of the DKT parcellation. These IDP sets were specifically compared in terms of the predictive capability vis-à-vis *Age* and *Genetic Sex*. With respect to UKBB-derived cortical thickness IDPs, similar analysis using both linear and XGBoost models demonstrates consistency with prior results (see Figure 4).

**Figure 4:**
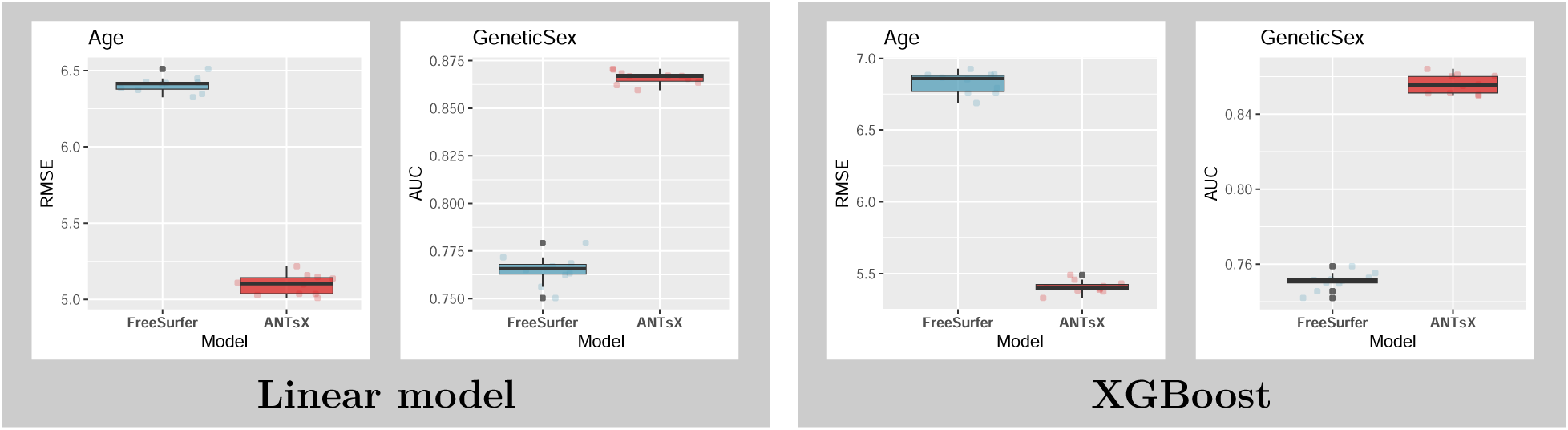
(Left) Linear and (right) XGBoost model results for predicting *Age* and *Genetic Sex* using both ANTsX and FreeSurfer cortical thickness data averaged over the 62 cortical regions of the DKT parcellation. RMSE and AUC were used to quantify the predictive accuracy of *Age* and *Genetic Sex*, respectively.

#### 3.1.2 Package IDP comparison via continuous target variables

Predictive models for cohort *Age*, *Fluid Intelligence Score*, *Neuroticism Score*, *Numeric Memory*, *Body Mass Index*, and *Townsend Deprivation Index* were generated and evaluated as described previously. Summary statistics for these variables are provided in Table 2. The resulting accuracies, in terms of RMSE, are provided in Figure 5.

**Figure 5:**
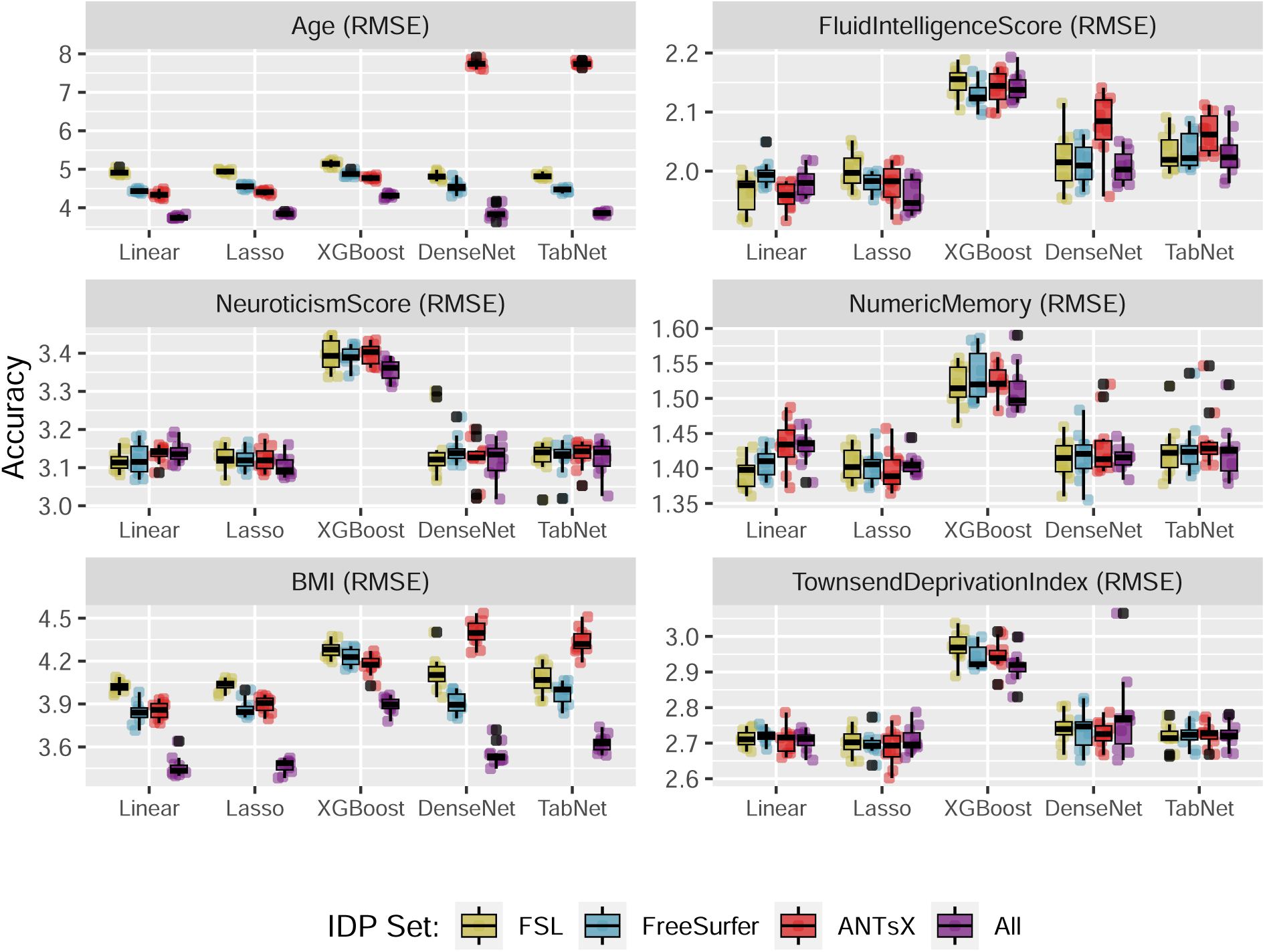
Comparison of machine learning frameworks for training and prediction of selected continuous UKBB sociodemographic continuous variables (cf. Table 1) with the different IDP sets and their combination (FSL, FreeSurfer, ANTsX, and All).

**Table 2:**
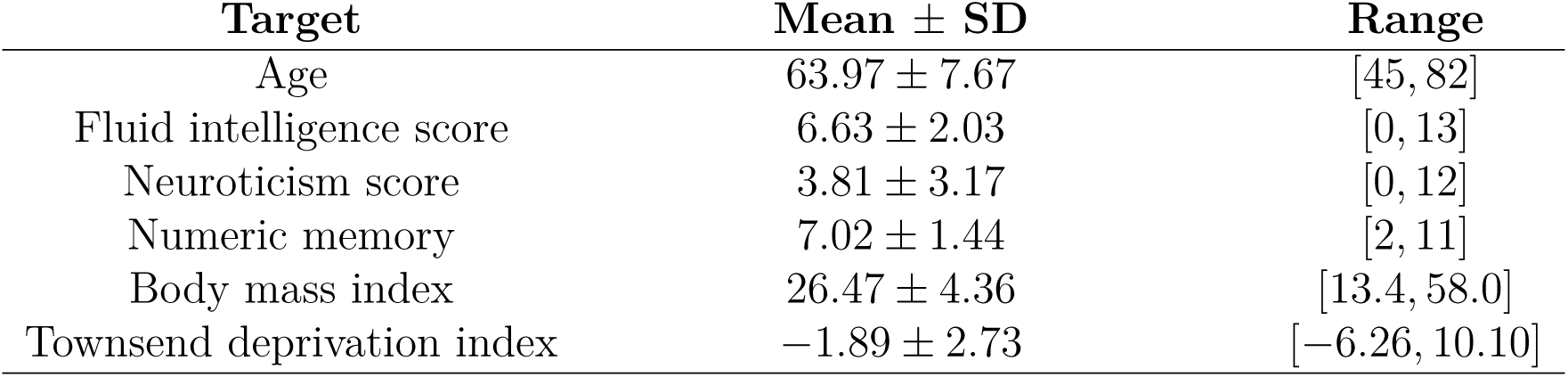
Summary statistics for the selected continuous UKBB sociodemographic target variables.

| Target | Mean ± SD | Range |
| --- | --- | --- |
| Age | $63.97 \pm 7.67$ | [45, 82] |
| Fluid intelligence score | $6.63 \pm 2.03$ | [0, 13] |
| Neuroticism score | $3.81 \pm 3.17$ | [0, 12] |
| Numeric memory | $7.02 \pm 1.44$ | [2, 11] |
| Body mass index | $26.47 \pm 4.36$ | [13.4, 58.0] |
| Townsend deprivation index | $-1.89 \pm 2.73$ | [-6.26, 10.10] |

Linear and Lasso models provide the most consistently accurate results across the set of continuous target variables with the combined set of IDPs performing well for the majority of cases. All linear models demonstrate significant correlations across IDP sets (cf. Figure 6). Despite the large number of regressors, sparsity-based constraints did not significantly improve prediction performance. The deep learning models (both DenseNet and TabNet) performed similarly although were only competitive for selected subsets (e.g., *Neuroticism Score* and *Townsend Deprivation Index*). Although XGBoost performed well for the commonly studied *Age* target variable, performance measures were relatively much less accurate for the remaining categories. This could be the results of suboptimal hyperparameter choice with respect to these other categories but, as with the other techniques, this was not investigated further.

**Figure 6:**
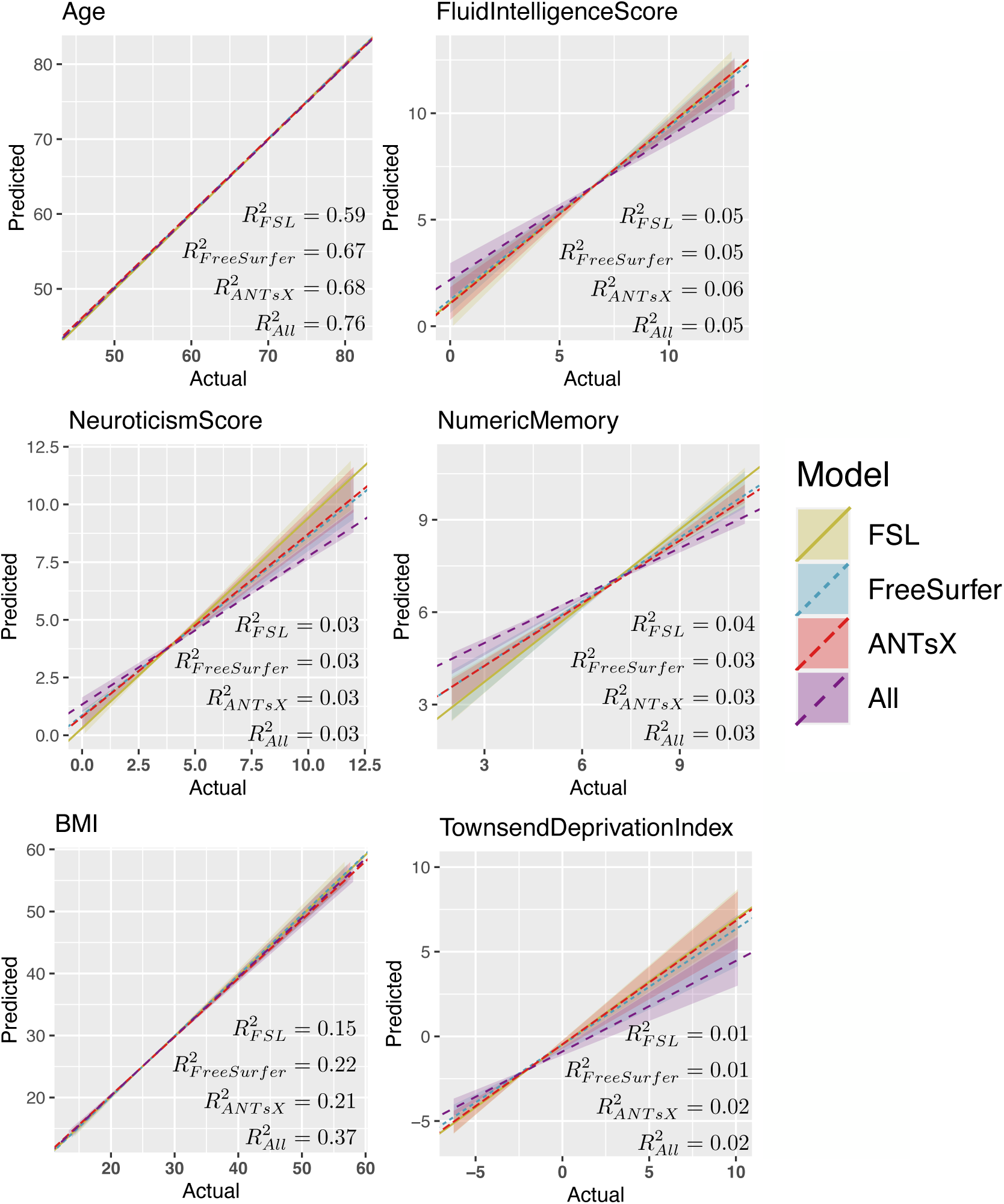
Regression regions defined by the linear models represented in Figure 5 showing the relationship between the predicted and actual target values. We also plot the median line for each model-based grouping as defined by the slope and list the average *R*^2^ values for each IDP set.

#### 3.1.3 Package IDP comparison via categorical target variables

Predictive models for cohort categories associated with *Genetic Sex*, *Hearing Difficulty*, *Risk Taking*, *Same Sex Intercourse*, *Smoking Frequency*, and *Alcohol Frequency* were generated and evaluated as described previously. The resulting accuracies, in terms of binary or multiclass AUC, are provided in Figure 7.

**Figure 7:**
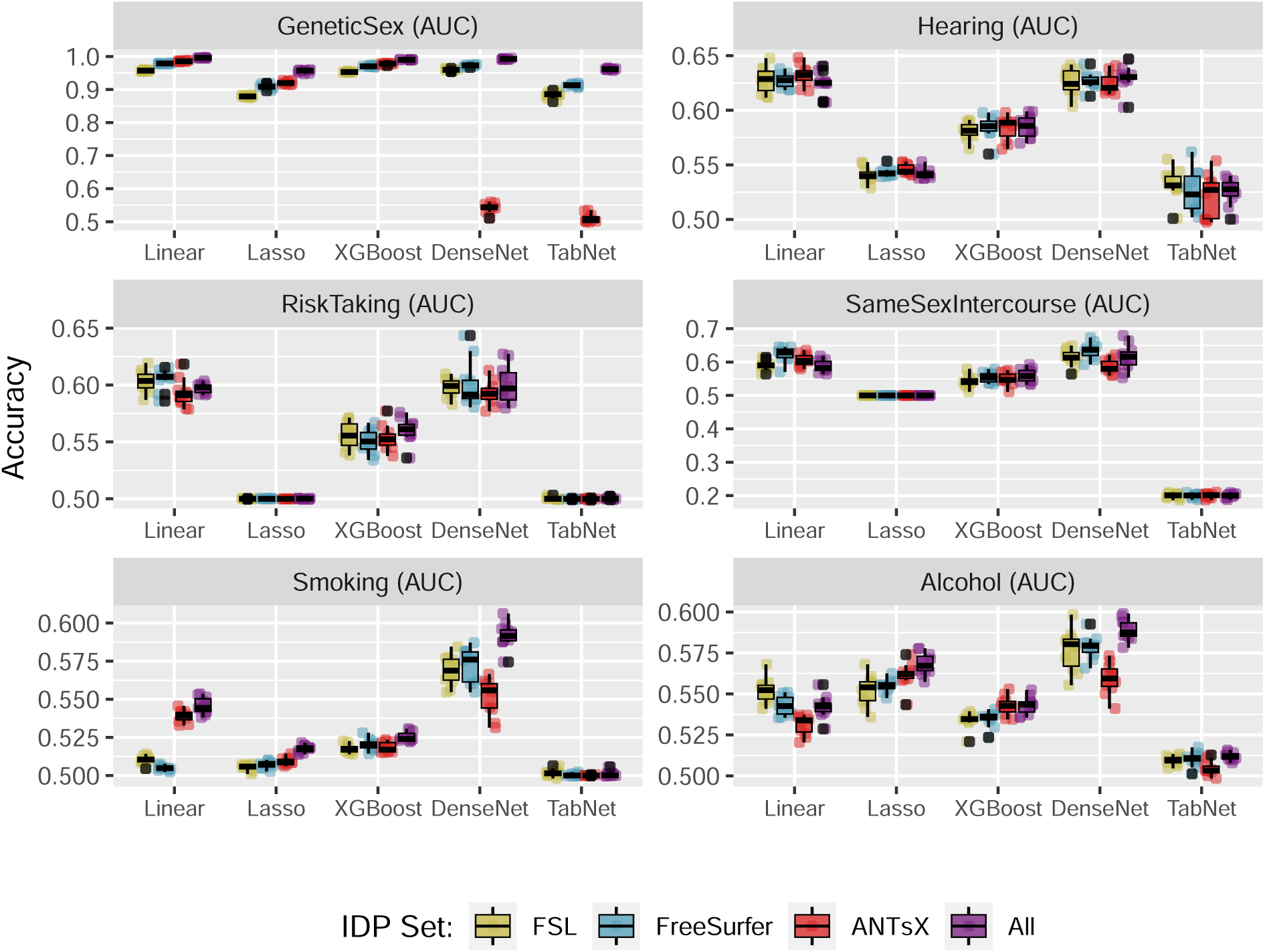
Comparison of machine learning frameworks for training and prediction of selected binary and multilabel categorical UKBB sociodemographic variables (cf. Table 1) with the different IDP sets and their combination (FSL, FreeSurfer, ANTsX, and All). *Smoking* and *Alcohol* target variables have more than two labels.

Similar to the continuous variables, the Linear and Lasso models perform well for most of the target variables. Superior performance is seen for these models predicting *Genetic Sex* along with XGBoost. DenseNet performs similarly well as the Linear and Lasso models for *Hearing*, *Risk Taking*, *Same Sex Intercourse*, and *Alcohol*. DenseNet models are also superior performers for predicting *Smoking* categories.

### 3.2 Individual IDP comparison

To compare individual IDPs, for each target variable, we selected the set of results corresponding to the machine learning technique which demonstrated superior performance, in terms of median predictive accuracy, for the combined (All) IDP grouping. The top ten features for the principle continuous variables of *Age*, *Fluid Intelligence Score*, and *Neuroticism Score* are listed in Table 3 and ranked according to variable importance score (specifically, absolute t-statistic value for linear models). The ranked lists are also color-coded by IDP package. For additional insight into individual IDPs, full feature lists with feature importance rankings are available for all target variables in the supplementary material hosted at the corresponding GitHub repository.^67^

**Table 3:**
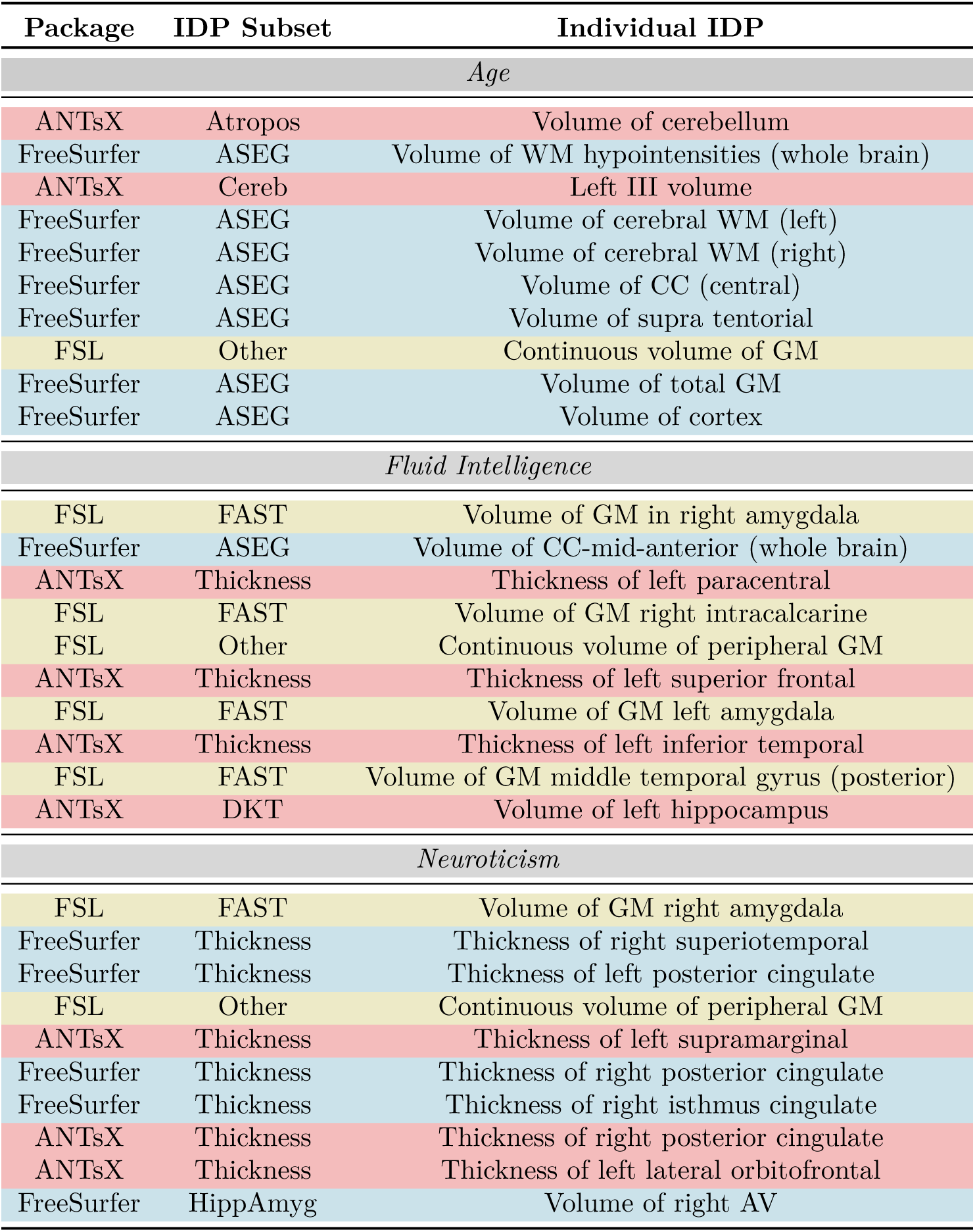
Top 10 features for *Age*, *Fluid Intelligence Score*, and *Neuroticism Score* target variables specified based on the specific, top-performing machine learning techniques for the combined (i.e., All) IDP set.

| Package | IDP Subset | Individual IDP |
| --- | --- | --- |
| *Age* |  |  |
| ANTsX | Atropos | Volume of cerebellum |
| FreeSurfer | ASEG | Volume of WM hypointensities (whole brain) |
| ANTsX | Cereb | Left III volume |
| FreeSurfer | ASEG | Volume of cerebral WM (left) |
| FreeSurfer | ASEG | Volume of cerebral WM (right) |
| FreeSurfer | ASEG | Volume of CC (central) |
| FreeSurfer | ASEG | Volume of supra tentorial |
| FSL | Other | Continuous volume of GM |
| FreeSurfer | ASEG | Volume of total GM |
| FreeSurfer | ASEG | Volume of cortex |
| *Fluid Intelligence* |  |  |
| FSL | FAST | Volume of GM in right amygdala |
| FreeSurfer | ASEG | Volume of CC-mid-anterior (whole brain) |
| ANTsX | Thickness | Thickness of left paracentral |
| FSL | FAST | Volume of GM right intracalcarine |
| FSL | Other | Continuous volume of peripheral GM |
| ANTsX | Thickness | Thickness of left superior frontal |
| FSL | FAST | Volume of GM left amygdala |
| ANTsX | Thickness | Thickness of left inferior temporal |
| FSL | FAST | Volume of GM middle temporal gyrus (posterior) |
| ANTsX | DKT | Volume of left hippocampus |
| *Neuroticism* |  |  |
| FSL | FAST | Volume of GM right amygdala |
| FreeSurfer | Thickness | Thickness of right superiotemporal |
| FreeSurfer | Thickness | Thickness of left posterior cingulate |
| FSL | Other | Continuous volume of peripheral GM |
| ANTsX | Thickness | Thickness of left supramarginal |
| FreeSurfer | Thickness | Thickness of right posterior cingulate |
| FreeSurfer | Thickness | Thickness of right isthmus cingulate |
| ANTsX | Thickness | Thickness of right posterior cingulate |
| ANTsX | Thickness | Thickness of left lateral orbitofrontal |
| FreeSurfer | HippAmyg | Volume of right AV |

## 4 Discussion

Much UKBB research is made possible through the availability of its characteristic largescale, subject-specific epidemiological data, including IDPs and enhanced by the stringent data acquisition protocols to ensure consistency across sites. In this work, we complement the existing FSL- and FreeSurfer-based UKBB IDPs with the generation and potential distribution of corresponding ANTsX-derived IDPs. These latter IDPs were generated from well-vetted pipelines that have been used in previous research and are publicly available through the ANTsX ecosystem. By providing these IDP-producing utilities within high-level languages, such as Python and R, in a comprehensive, open-source package, we are able to leverage the computational efficiency of deep learning libraries while also leveraging the numerous packages available for the curation, analysis, and visualization of tabulated data.

In addition to the availability of these ANTsX UKBB IDPs, we explored their utility with respect to other package-specific groupings and their combinations. For exploration of these IDP group permutations, we used popular machine learning algorithms to predict commonly studied sociodemographic variables of current research interest (Table 1). In addition to research presentation in traditional venues, at least two of these target variables, specifically *Age* and *Fluid Intelligence*, have been the focus of two recent competitions.

Regarding the former, research concerning brain age estimation from neuroimaging is extensive and growing (cf. recent reviews).^37,68,69^ It was also the subject of the recent Predictive Analytics Competition held in 2019 (PAC2019). This competition featured 79 teams leveraging T1-weighted MRI with a variety of quantitative approaches from convolutional neural networks (CNNs) to common machine learning frameworks based on morphological descriptors (i.e., structural IDPs) derived from FreeSurfer.^70^ The winning team,^71^ using an ensemble of CNNs and pretrained on a UKBB cohort of *N* = 14, 503 subjects, had a mean absolute error (MAE) of 2.90 years. Related CNN-based deep learning approaches achieved comparable performance levels and simultaneously outperformed more traditional machine learning approaches. For example, the FreeSurfer IDP approach using a dense neural network^70^ yielded an overall MAE accuracy of 4.6 years. Alternative strategies based on Lasso, Random Forests, and support vector regression techniques were attempted but did not achieve similar accuracy levels.

Given that RMSE provides a general upper bound on MAE (i.e., MAE *≤* RMSE), the accuracy levels yielded by our FSL, FreeSurfer, ANTsX, and combined linear, lasso, and XGBoost models can be seen from Figure 5 to perform comparatively well. The FreeSurfer and ANTsX linear models performed similarly with RMSE prediction values of approximately 4.4 years whereas FSL was a little higher at 4.96 years. However, combining all IDPs resulted in an average RMSE value of 3.8 years. When looking at the top 10 overall linear model features (Table 3) ranked in terms of absolute t-statistic value, all three packages are represented and appear to reflect both global structures (white matter and CSF volumes) and general subcortical structural volumes (ANTsX “deep GM” and both FreeSurfer and ANTsX bi-hemispherical ventral dienchephalon volumes). Increases in CSF volume and ventricular spaces is well-known to be associated with brain shrinkage and aging.^72–74^

Similarly, the association between brain structure and fluid intelligence has been well-studied^75^ despite potentially problematic philosophical and ethical issues.^76^ With intentions of furthering this research, the ABCD Neurocognitive Prediction Challenge (ABCD-NP- Challenge) was held in 2019 which concerned predicting fluid intelligence scores (using the NIH Toolbox Cognition Battery)^77^ in a population of 9-10 year pediatric subjects using T1-weighted MRI. Fluid intelligence scores were residualized from brain volume, acquisition site, age, ethnicity, genetic sex, and parental attributes of income, education, and marriage (additional data processing details are provided in the Data Supplement).^78^

Of the 29 submitting teams, the first place team of the final leaderboard employed kernel ridge regression with voxelwise features based on the T1-weighted-based probabilistic tissue segmentations specifically, CSF, gray matter, and white matter— both modulated and unmodulated versions for a total of six features per subject. In contrast to the winning set of predictive sparse and global features, the second place team used 332 total cortical, subcortical, white matter, cerebellar, and CSF volumetric features. Although exploring several machine learning modeling techniques, the authors ultimately used an ensemble of models for prediction which showed improvement over gradient boosted decision trees. From Table 3, most predictive features from our study, regardless of package, are localized measures of gray matter.

Although the stated, primary objective of these competitions is related to superior performance in terms of algorithmic prediction of quantitative sociodemographics, similar to the evaluation strategy used in this work, outside of the clinical research into brain age estimation, none of these performance metric reach the level of individual-level prediction. Consequently, these may be more informative as an interpretation of the systems- level relationship between brain structure and behavior. An obvious secondary benefit is the insight gained into the quality and relevance of measurements and modeling techniques used. In this way, these considerations touch on fundamental implications of the No Free Lunch Theorems for search and optimization^79^ where prior distributions (i.e., correspondence of measurements and clinical domain for algorithmic modeling) differentiate general performance. Relatedly, although all packages are represented amongst the top-performing IDPs, their relative utility is dependent, expectedly so, on the specific target variable, and, to a lesser extent, on the chosen machine learning technique. Such considerations should be made along with other relevant factors (e.g., computational requirements, open-source availability) for tailored usage.

## 5 Conclusion

The UK Biobank is an invaluable resource for large-scale epidemiological research which includes a thorough neuroimaging battery for a significant subset of the study volunteers. For quantitative exploration and inference of population trends from leveraging imaging data, well-vetted measurement tools are essential. The primary contribution that we have described is the generation and public availability of the set of UK Biobank neuroimaging structural IDPs generated using the ANTsX ecosystem. These ANTsX IDPs, which includes DeepFLASH for hippocampal and extra-hippocampal parcellation, complement the existing sets of FSL and FreeSurfer IDPs. A predictive modeling strategy using a variety of sociodemographic target variables was used to explore IDP viability, importance, and utility via the modeling capabilities of several machine learning techniques with linear regression demonstrating overall good performance.

## Data Availability

All tabular data and analysis scripts described in the paper (including the source files for the paper) are available online at https://github.com/ntustison/ANTsXUKBB.

https://github.com/ANTsX

## Footnotes

1 “Observer bias” is considered in the context of casting computational measurement tools as “observers” with “observer bias” due to the specific set of choices that results in the final numerical measurement. These choices can include (but are certainly not limited to) modeling considerations, preferences with respect to anatomical definitional ambiguities, and the set of parameters used to run the corresponding software.

## References

1. Cornelis, M. C. et al. Age and cognitive decline in the UK biobank. PLoS One 14, e0213948 (2019).

2. Hagenaars, S. P. et al. Shared genetic aetiology between cognitive functions and physical and mental health in UK biobank (n=112,151) and 24 GWAS consortia. Mol Psychiatry 21, 1624–1632 (2016).

3. Griffith, G. J. et al. Collider bias undermines our understanding of COVID-19 disease risk and severity. Nat Commun 11, 5749 (2020).

4. Miller, K. L. et al. Multimodal population brain imaging in the UK biobank prospective epidemiological study. Nat Neurosci 19, 1523–1536 (2016).

5. Alfaro-Almagro, F. et al. Image processing and quality control for the first 10,000 brain imaging datasets from UK biobank. Neuroimage 166, 400–424 (2018).

6. Nobis, L. et al. Hippocampal volume across age: Nomograms derived from over 19,700 people in UK biobank. Neuroimage Clin 23, 101904 (2019).

7. Dadi, K. et al. Population modeling with machine learning can enhance measures of mental health. Gigascience 10, (2021).

8. Douaud, G. et al. SARS-CoV-2 is associated with changes in brain structure in UK biobank. Nature (2022) doi:10.1038/s41586-022-04569-5.

9. Jenkinson, M., Beckmann, C. F., Behrens, T. E. J., Woolrich, M. W. & Smith, S. M. FSL. Neuroimage 62, 782–90 (2012).

10. Zhang, Y., Brady, M. & Smith, S. Segmentation of brain MR images through a hidden markov random field model and the expectation-maximization algorithm. IEEE Trans Med Imaging 20, 45–57 (2001).

11. Patenaude, B., Smith, S. M., Kennedy, D. N. & Jenkinson, M. A bayesian model of shape and appearance for subcortical brain segmentation. Neuroimage 56, 907–22 (2011).

12. Griffanti, L. et al. BIANCA (brain intensity AbNormality classification algorithm): A new tool for automated segmentation of white matter hyperintensities. Neuroimage 141, 191–205 (2016).

13. Fischl, B. FreeSurfer. Neuroimage 62, 774–81 (2012).

14. Iglesias, J. E. et al. A computational atlas of the hippocampal formation using ex vivo, ultra-high resolution MRI: Application to adaptive segmentation of in vivo MRI. Neuroimage 115, 117–37 (2015).

15. Saygin, Z. M. et al. High-resolution magnetic resonance imaging reveals nuclei of the human amygdala: Manual segmentation to automatic atlas. Neuroimage 155, 370–382 (2017).

16. Tustison, N. J. et al. The ANTsX ecosystem for quantitative biological and medical imaging. Sci Rep 11, 9068 (2021).

17. Avants, B. B. et al. A reproducible evaluation of ANTs similarity metric performance in brain image registration. Neuroimage 54, 2033–44 (2011).

18. Avants, B. B. et al. The Insight ToolKit image registration framework. Front Neuroinform 8, 44 (2014).

19. Yoo, T. S. & Metaxas, D. N. Open science–combining open data and open source software: Medical image analysis with the insight toolkit. Med Image Anal 9, 503–6 (2005).

20. Ding, A. S. et al. Automated extraction of anatomical measurements from temporal bone CT imaging. Otolaryngol Head Neck Surg 1945998221076801 (2022) doi:10.1177/01945998221076801.

21. Diamond, K. M., Rolfe, S. M., Kwon, R. Y. & Maga, A. M. Computational anatomy and geometric shape analysis enables analysis of complex craniofacial phenotypes in zebrafish. Biol Open 11, (2022).

22. Kini, L. G. et al. Quantitative [18]FDG PET asymmetry features predict long-term seizure recurrence in refractory epilepsy. Epilepsy Behav 116, 107714 (2021).

23. Avants, B. B., Tustison, N. J., Wu, J., Cook, P. A. & Gee, J. C. An open source multivariate framework for n-tissue segmentation with evaluation on public data. Neuroinformatics 9, 381–400 (2011).

24. Tustison, N. J. et al. Large-scale evaluation of ANTs and FreeSurfer cortical thickness measurements. Neuroimage 99, 166–79 (2014).

25. Tustison, N. J. et al. Longitudinal mapping of cortical thickness measurements: An Alzheimer’s Disease Neuroimaging Initiative-based evaluation study. J Alzheimers Dis (2019) doi:10.3233/JAD-190283.

26. Klein, A. & Tourville, J. 101 labeled brain images and a consistent human cortical labeling protocol. Front Neurosci 6, 171 (2012).

27. Reagh, Z. M. et al. Functional imbalance of anterolateral entorhinal cortex and hippocampal dentate/CA3 underlies age-related object pattern separation deficits. Neuron 97, 1187–1198.e4 (2018).

28. Schmahmann, J. D. et al. Three-dimensional MRI atlas of the human cerebellum in proportional stereotaxic space. Neuroimage 10, 233–60 (1999).

29. Das, S. R., Avants, B. B., Grossman, M. & Gee, J. C. Registration based cortical thickness measurement. Neuroimage 45, 867–79 (2009).

30. Weiner, M. W. et al. The alzheimer’s disease neuroimaging initiative: A review of papers published since its inception. Alzheimers Dement 8, S1–68 (2012).

31. Caliskan, A., Bryson, J. J. & Narayanan, A. Semantics derived automatically from language corpora contain human-like biases. Science 356, 183–186 (2017).

32. Tustison, N. J. et al. Instrumentation bias in the use and evaluation of scientific software: Recommendations for reproducible practices in the computational sciences. Front Neurosci 7, 162 (2013).

33. Yushkevich, P. A. et al. Quantitative comparison of 21 protocols for labeling hippocampal subfields and parahippocampal subregions in in vivo MRI: Towards a harmonized segmentation protocol. Neuroimage 111, 526–41 (2015).

34. Chen, T. & Guestrin, C. XGBoost: A scalable tree boosting system. in Proceedings of the 22nd ACM SIGKDD international conference on knowledge discovery and data mining 785–794 (ACM, 2016). doi:10.1145/2939672.2939785.

35. Arik, S. Ö. & Pfister, T. TabNet: Attentive interpretable tabular learning. Proceedings of the AAAI Conference on Artificial Intelligence 35, 6679–6687 (2021).

36. Lundberg, S. M. & Lee, S.-I. A unified approach to interpreting model predictions. in Advances in neural information processing systems 30 (eds. Guyon, I. et al.) 4765–4774 (Curran Associates, Inc., 2017).

37. Franke, K. & Gaser, C. Ten years of BrainAGE as a neuroimaging biomarker of brain aging: What insights have we gained? Front Neurol 10, 789 (2019).

38. Nyberg, L. & Wåhlin, A. The many facets of brain aging. Elife 9, (2020).

39. Smith, S. M. et al. Brain aging comprises many modes of structural and functional change with distinct genetic and biophysical associations. Elife 9, (2020).

40. UKBB.

41. Glasser, M. F. et al. The minimal preprocessing pipelines for the human connectome project. Neuroimage 80, 105–24 (2013).

42. Falk, T. et al. U-net: Deep learning for cell counting, detection, and morphometry. Nat Methods 16, 67–70 (2019).

43. Fonov, V. S., Evans, A. C., McKinstry, R. C., Almli, C. & Collins, D. L. Unbiased nonlinear average age-appropriate brain templates from birth to adulthood. NeuroImage **S102**, (2009).

44. Schlemper, J. et al. Attention gated networks: Learning to leverage salient regions in medical images. Med Image Anal 53, 197–207 (2019).

45. Desikan, R. S. et al. An automated labeling system for subdividing the human cerebral cortex on MRI scans into gyral based regions of interest. Neuroimage 31, 968–80 (2006).

46. Sethian, J. A. A fast marching level set method for monotonically advancing fronts. Proc Natl Acad Sci U S A 93, 1591–5 (1996).

47. Wang, H. et al. Multi-atlas segmentation with joint label fusion. IEEE Trans Pattern Anal Mach Intell 35, 611–23 (2013).

48. Brown, E. S. et al. A randomized, double-blind, placebo-controlled trial of lamotrigine for prescription corticosteroid effects on the human hippocampus. Eur Neuropsychopharmacol 29, 376–383 (2019).

49. Brown, E. S. et al. A randomized trial of an NMDA receptor antagonist for reversing corticosteroid effects on the human hippocampus. Neuropsychopharmacology 44, 2263–2267 (2019).

50. Holbrook, A. J. et al. Anterolateral entorhinal cortex thickness as a new biomarker for early detection of alzheimer’s disease. Alzheimers Dement (Amst) 12, e12068 (2020).

51. McMakin, D. L., Kimbler, A., Tustison, N. J., Pettit, J. W. & Mattfeld, A. T. Negative overgeneralization is associated with anxiety and mechanisms of pattern completion in peripubertal youth. Soc Cogn Affect Neurosci (2021) doi:10.1093/scan/nsab089.

52. Nguyen, D. M. et al. The relationship between cumulative exogenous corticosteroid exposure and volumes of hippocampal subfields and surrounding structures. J Clin Psychopharmacol 39, 653–657 (2019).

53. Sinha, N. et al. APOE *ε*4 status in healthy older african americans is associated with deficits in pattern separation and hippocampal hyperactivation. Neurobiol Aging 69, 221–229 (2018).

54. Tustison, N. J. & Avants, B. B. ANTsRNet GitHub. https://github.com/ANTsX/ANTsRNet.

55. Tustison, N. J. & Avants, B. B. ANTsPyNet GitHub. https://github.com/ANTsX/ANTsPyNet.

56. Park, M. T. M. et al. Derivation of high-resolution MRI atlases of the human cerebellum at 3T and segmentation using multiple automatically generated templates. Neuroimage 95, 217–31 (2014).

57. Li, H. et al. Fully convolutional network ensembles for white matter hyperintensities segmentation in MR images. Neuroimage 183, 650–665 (2018).

58. Kuijf, H. J. et al. Standardized assessment of automatic segmentation of white matter hyperintensities and results of the WMH segmentation challenge. IEEE Trans Med Imaging 38, 2556–2568 (2019).

59. Kuhn, M. Building predictive models in r using the caret package. Journal of Statistical Software 28, 1–26 (2008).

60. Friedman, J., Hastie, T. & Tibshirani, R. Regularization paths for generalized linear models via coordinate descent. Journal of Statistical Software 33, 1–22 (2010).

61. Breiman, L. Random forests. Machine Learning 45, 5–32 (2001).

62. Friedman, J. H. Greedy function approximation: A gradient boosting machine. The Annals of Statistics 29, 1189–1232 (2001).

63. McTabberson, T. https://github.com/dreamquark-ai/tabnet.

64. Shwartz-Ziv, R. & Armon, A. Tabular data: Deep learning is not all you need. Information Fusion 81, 84–90 (2022).

65. Kadra, A., Lindauer, M., Hutter, F. & Grabocka, J. Regularization is all you need: Simple neural nets can excel on tabular data. CoRR abs/2106.11189, (2021).

66. Gorishniy, Y., Rubachev, I., Khrulkov, V. & Babenko, A. Revisiting deep learning models for tabular data. CoRR abs/2106.11959, (2021).

67. Tustison, N. J. ANTsXUKBB GitHub. https://github.com/ntustison/ANTsXUKBB.

68. Mishra, S., Beheshti, I. & Khanna, P. A review of neuroimaging-driven brain age estimation for identification of brain disorders and health conditions. IEEE Rev Biomed Eng PP, (2021).

69. Baecker, L., Garcia-Dias, R., Vieira, S., Scarpazza, C. & Mechelli, A. Machine learning for brain age prediction: Introduction to methods and clinical applications. EBioMedicine 72, 103600 (2021).

70. Lombardi, A. et al. Brain age prediction with morphological features using deep neural networks: Results from predictive analytic competition 2019. Front Psychiatry 11, 619629 (2020).

71. Gong, W., Beckmann, C. F., Vedaldi, A., Smith, S. M. & Peng, H. Optimising a simple fully convolutional network for accurate brain age prediction in the PAC 2019 challenge. Front Psychiatry 12, 627996 (2021).

72. Murphy, D. G., DeCarli, C., Schapiro, M. B., Rapoport, S. I. & Horwitz, B. Agerelated differences in volumes of subcortical nuclei, brain matter, and cerebrospinal fluid in healthy men as measured with magnetic resonance imaging. Arch Neurol 49, 839–45 (1992).

73. Matsumae, M. et al. Age-related changes in intracranial compartment volumes in normal adults assessed by magnetic resonance imaging. J Neurosurg 84, 982–91 (1996).

74. Scahill, R. I. et al. A longitudinal study of brain volume changes in normal aging using serial registered magnetic resonance imaging. Arch Neurol 60, 989–94 (2003).

75. Vieira, B. H. et al. On the prediction of human intelligence from neuroimaging: A systematic review of methods and reporting. Intelligence 93, 101654 (2022).

76. Eickhoff, S. B. & Langner, R. Neuroimaging-based prediction of mental traits: Road to utopia or orwell? PLoS Biol 17, e3000497 (2019).

77. Weintraub, S. et al. Cognition assessment using the NIH toolbox. Neurology 80, S54–64 (2013).

78. Pfefferbaum, A. et al. Altered brain developmental trajectories in adolescents after initiating drinking. Am J Psychiatry 175, 370–380 (2018).

79. Wolpert, D. H. & Macready, W. G. No free lunch theorems for optimization. IEEE Transactions on Evolutionary Computation 1, 67–82 (1997).

